# Health Data Nexus: An Open Data Platform for AI Research and Education in Medicine

**DOI:** 10.1101/2024.08.23.24312060

**Authors:** January Adams, Rafal Cymerys, Karol Szuster, Daniel Hekman, Zoryana Salo, Rutvik Solanki, Muhammad Mamdani, Alistair Johnson, Katarzyna Ryniak, Tom Pollard, David Rotenberg, Benjamin Haibe-Kains

## Abstract

We outline the development of the Health Data Nexus, a data platform which enables data storage and access management with a cloud-based computational environment. We describe the importance of this secure platform in an evolving public sector research landscape that utilizes significant quantities of data, particularly clinical data acquired from health systems, as well as the importance of providing meaningful benefits for three targeted user groups: data providers, researchers, and educators. We then describe the implementation of governance practices, technical standards, and data security and privacy protections needed to build this platform, as well as example use-cases highlighting the strengths of the platform in facilitating dataset acquisition, novel research, and hosting educational courses, workshops, and datathons. Finally, we discuss the key principles that informed the platform’s development, highlighting the importance of flexible uses, collaborative development, and open-source science.

## BACKGROUND

The fast-growing field of machine learning (ML) and artificial intelligence (AI) holds transformative promise for the field of medicine, offering prospects in enhancing patient outcomes and improving healthcare delivery. These technologies are pivotal in driving advancements across a spectrum of applications, including the early detection of emergent medical conditions, the development of diagnostic and prognostic tests, and the acceleration of drug discovery and development.

The transformative potential of AI in healthcare depends on the availability and accessibility of large, high-quality datasets. The healthcare sector is a prolific generator of data, yet this information is often siloed, dispersed across various entities, and mired in issues of accessibility for research purposes. Moreover, the data landscape in healthcare is characterized by a lack of uniform standards [1], compounded by inadequate documentation, which collectively pose significant challenges to the effective use of this data in ML/AI applications.

To harness the full transformative potential of ML/AI in medicine, these barriers must be addressed, fostering an environment where data sharing is streamlined, standardized, and conducted in a manner that upholds the high standards of data privacy and security. Both comprehensive, well-curated datasets and advanced ML/AI methodologies are crucial to unlock innovative healthcare solutions that are precise, predictive, and patient-centric.

While many healthcare and health research organizations house data for secondary uses such as research, access to these datasets can be cumbersome as they are usually secured and only made available to researchers upon approval of a research plan and in collaboration with representatives of the organization. Once access is approved, researchers are often required to use their own external computing resources to analyze the data. While this process minimizes the risk of data misuse and leakage, it is lengthy and presents a significant barrier to health innovation through research.

Other data platforms have taken a more accessible approach to data sharing. One example is PhysioNet [2], a data sharing platform created and maintained by the Laboratory for Computational Physiology at the Massachusetts Institute of Technology in the US. PhysioNet provides a platform for users to share datasets and software and includes a workflow for approving and credentialing potential data users, who can then download data to work on their own projects. The entire platform is built as an open source project, allowing other researchers and institutions to make use of the same framework. This platform exists within an American regulatory environment where different restrictions apply, potentially limiting the access to some datasets. Other platforms such as Nightingale Open Science [3] use a similar framework for development.

Given the complexity and sensitivity of health data, striking the right balance between openness, ease-of-use, scalability and security remains an open challenge. Openness and ease-of-use are crucial to ensure that data can be accessed in an equitable and inclusive manner for a diverse group of users. As new technologies such as electronic medical records and sequencing are adopted, the amount and complexity of health data increases, calling for the design of digital platforms that are scalable from the perspective of computational resources. Further, many health datasets are highly sensitive and may remain so after de-identification, requiring the data platform to be secure and able to accommodate the potential requirements for access restrictions. Such restrictions can take the form of an approval from an Institutional Review Board (IRB) or Research Ethics Board (REB), a data usage agreement, or a review through a data access committee. Recognizing that generating high-quality health data requires expertise and resources, it is also essential that a platform offer strong incentives for data generators to contribute their datasets, creating a virtuous cycle between users and data providers.

To strike a balance between data security, usability, and streamlined access, we developed the Health Data Nexus (HDN) [4] within the Temerty Centre for Artificial Intelligence Research and Education in Medicine (T-CAIREM) [5]. T-CAIREM is a research centre at the University of Toronto. The HDN was developed as a platform for both the storage and analysis of de-identified health data for the academic research and education communities in Canada. Building upon the scalable Google Cloud Platform (GCP) and PhysioNet’s user-friendly interface, we have implemented multiple options for data access and use. This makes HDN an open-source and well-balanced platform for research and education. Here, we describe key characteristics of the HDN platform, its implementation, and its usage to transform research and education for AI in medicine.

## MOTIVATION

Acquiring real-world health data traditionally involves hosting data directly from healthcare systems; however, this approach frequently encounters significant barriers due to data security and privacy. Consequently, data providers must impose stringent restrictions on data availability and implement rigorous limitations on access, often complicating and slowing research initiatives. An alternative solution involves the use of synthetic data, though this approach introduces its own set of challenges, notably in preserving data realism and ensuring applicability of analytical findings to actual clinical scenarios. Recognizing these complexities, we developed the Health Data Nexus (HDN) at the intersection of data security, accessibility, and usability with three principal objectives—dataset curation, research enablement, and educational support—each tailored to meet distinct user group requirements (Figure 1). Below, we describe these core goals, the specific user groups they represent, and the technical and governance challenges addressed to achieve them.

**Figure 1.**
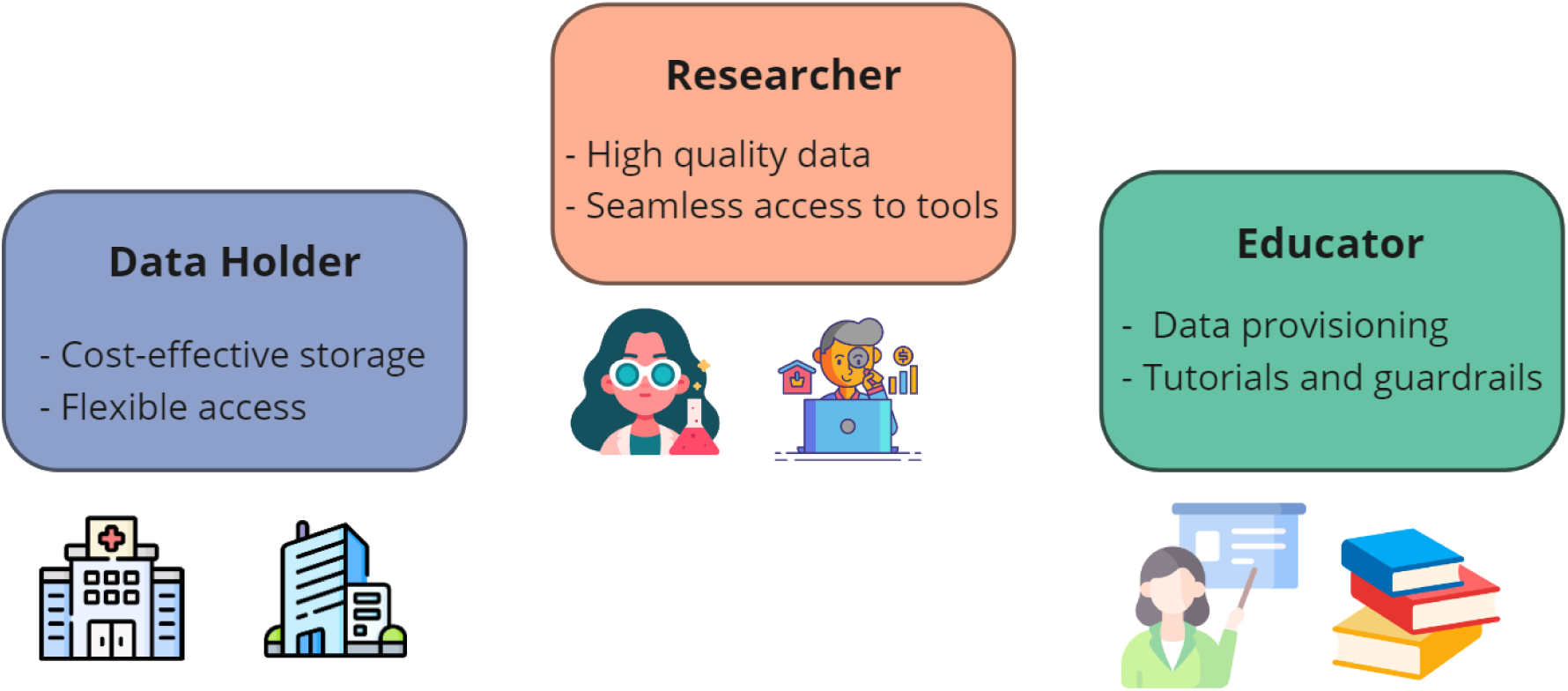
Summary of the three main user groups and their requirements on the platform.

### Data Holders

Data Holders seek simple, cost-effective, and secure data storage tailored to their individual needs. Whether they aim to make their data broadly available to a large number of potential users or only to a single lab or small group of trusted researchers, this requires a streamlined process. This process should include simple and understandable rules around governance and data hosting, as well as easy access to a Data Steward to assist with the process. Providers may also need flexibility around approving different levels of access to various potential users, allowing them to maintain control over their data while accommodating the needs of researchers.

### Researchers

Researchers require access to high quality data, along with the tools and support needed to conduct their analyses. They require the ability to seamlessly load data into a safe and secure research environment equipped with packages, essential software, and computing resources. Researchers also benefit from access to learning resources, such as user-contributed tutorials and methods or comprehensive metadata to describe the dataset. Helpful built-in support mechanisms can enable higher quality and timely completion of their projects; examples of these supports include access to knowledgeable support staff or a community of researchers to resolve research questions.

### Educators

Educators seek to distribute data using the platform, either to their students or to the attendees of a workshop or datathon. Educators benefit from a simple and streamlined method to provide access to datasets, complemented by tutorials that can be integrated into data environments as a starting point. They focus on provisioning data access to many users, emphasizing simplicity and ease over powerful computing resources. Educators may also benefit from mechanisms to prevent students or trainees from using more computing resources than required, preventing users from incurring greater financial costs.

## IMPLEMENTATION

Based on the user-specific needs, we outline the requirements for the HDN platform, including both governance requirements for data access (Figure 2) and technical requirements around data storage and the associated analysis platform (Figure 3).

**Figure 2.**
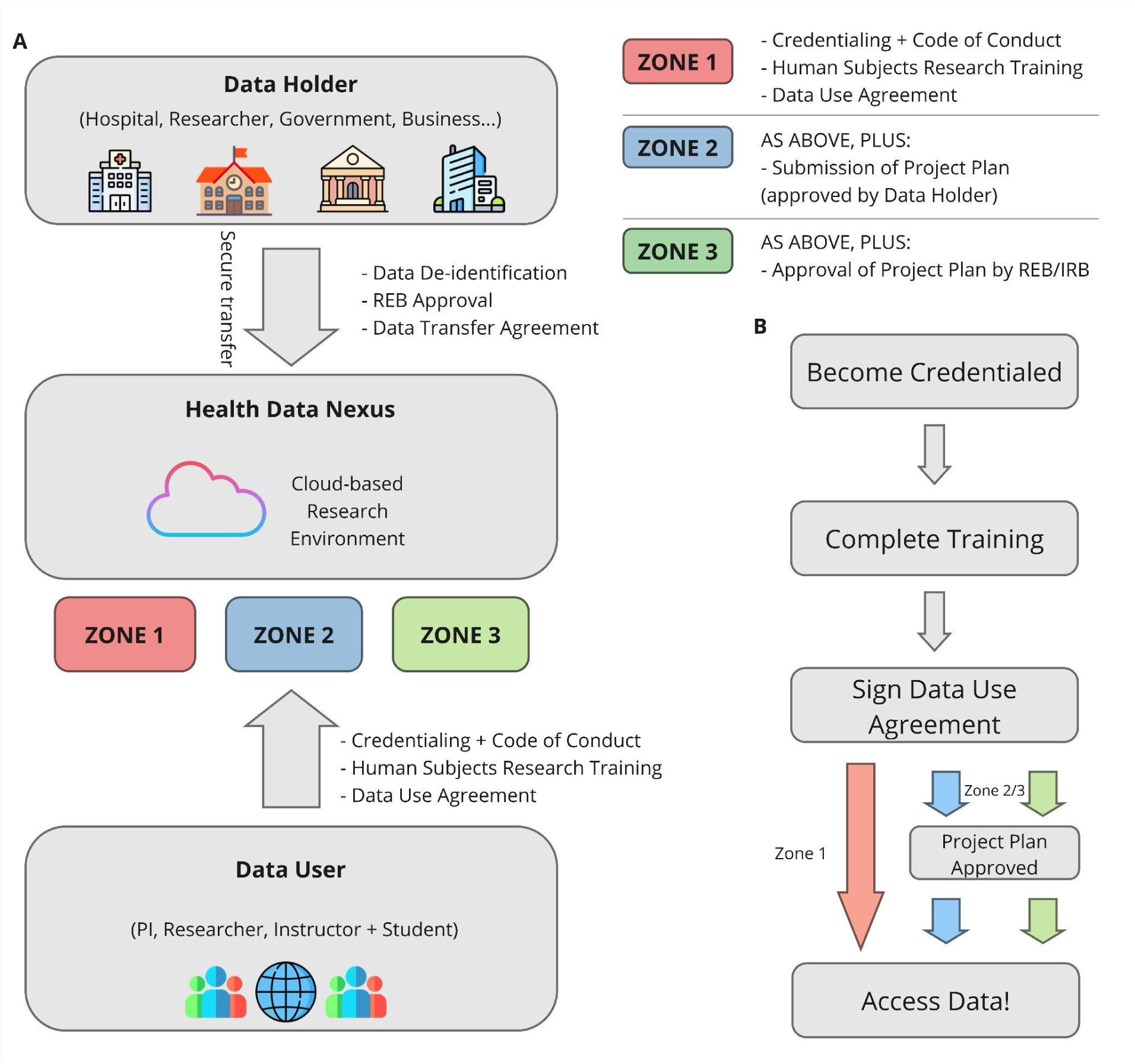
(A) An overview of the workflow for uploading data to the platform (for data holders) and accessing data (for data users), highlighting the different zones of access. (B) Details of the process for users to access data.

**Figure 3.**
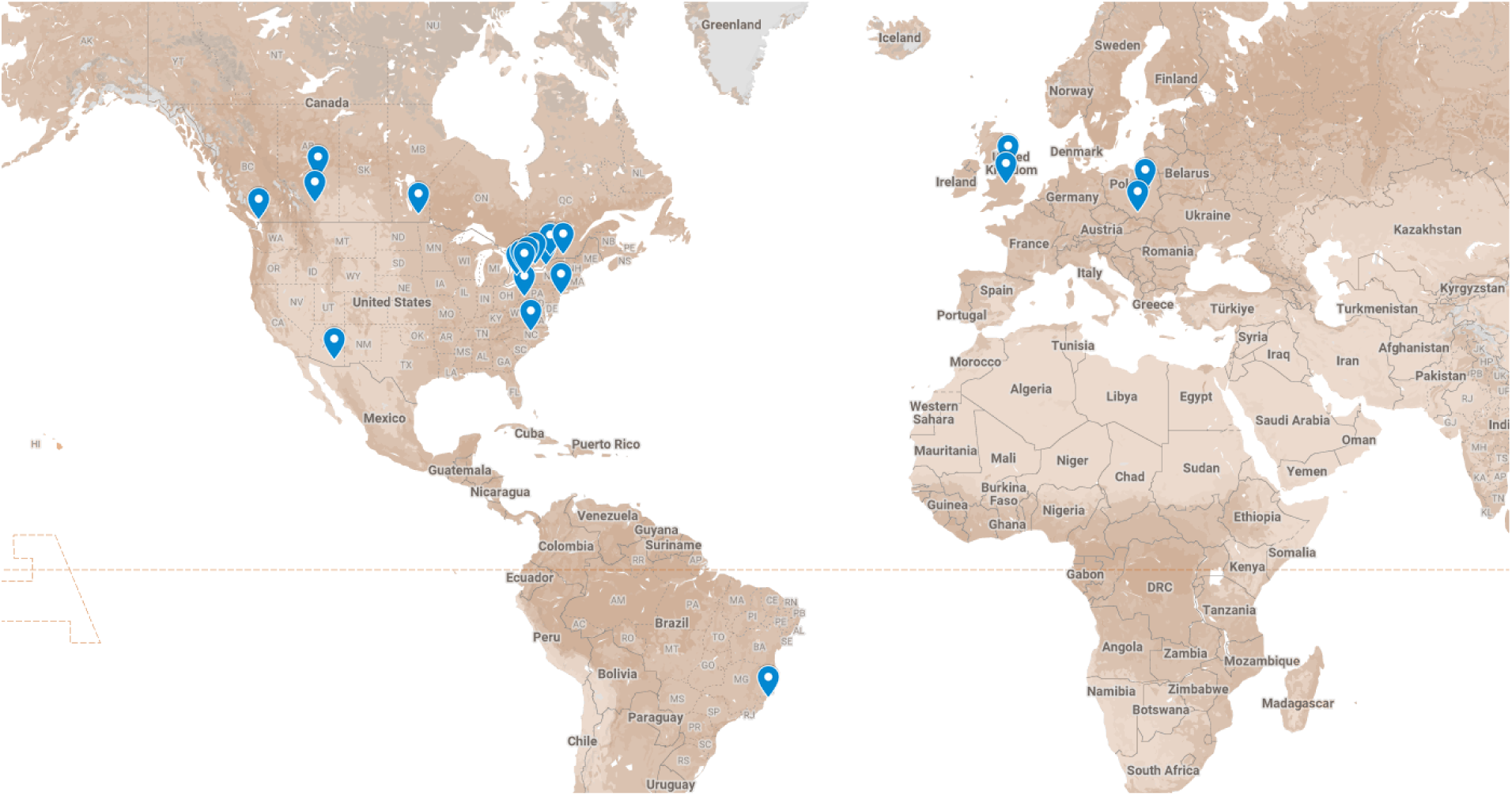
Map of universities in which at least one user with an associated university email address has accessed the platform.

### User Interface

The user-facing interface of the HDN platform has been designed to meet the needs of its user groups by being specific, social, and streamlined. We define a platform as specific if it is designed to meet the direct needs of users–in our case, the platform should be focused on health data, and use familiar tools and resources to do so. The frontend of the HDN platform is based on the PhysioNet platform, which provides the credentialing model and brings the capability to manage datasets, data users, and their access to the datasets. PhysioNet’s interface was chosen partially due to its familiarity to users, as many health researchers will be familiar with the interface required to access the very popular MIMIC-IV dataset. In addition, the PhysioNet structure was designed to handle multimodal data, including electrophysiological signals, images, tabular data, and free text, including combinations of these sources within a single dataset.

We also designed the user interface to be social, primarily in that it is open source and facilitates collaboration within the wider scientific community. The source code of PhysioNet, which underpins the Health Data Nexus, is accessible under the BSD 3-Clause license. During development of the Health Data Nexus, its code underwent significant modifications to accommodate the hosting of various data repositories across different scientific domains. We adjusted the platform to follow 12-factor app principles [6], allowing it to be deployed via the Google Kubernetes Engine, used Google Cloud Storage for storing datasets, and introduced a fully automated Continuous Integration and Continuous Delivery pipeline. These adjustments all update the Health Data Nexus to adapt to a modern technical infrastructure. These features were implemented back into the PhysioNet repository, allowing others to make use of them in the future and furthering our commitment to development through open science.

Finally, we aim for the user interface to provide a simple approach to managing dataset access. The Health Data Nexus supports curation and review processes for dataset publication and leverages Google Cloud Storage for internal data storage and managing dataset access permissions on the cloud-level. Additionally, the interface facilitates user access management, overseeing the credentialing process and setting access prerequisites for datasets, such as mandatory training completion. To expedite credentialing, the platform enables account creation via eduGAIN sign-in, which verifies university affiliations. Moreover, the frontend supports event management, particularly for datathons, by offering features such as temporary dataset access. This functionality is crucial for ensuring data use compliance during specific events.

### Computational Infrastructure

The computational infrastructure of the platform has been designed to be secure, scalable, and streamlined. Security on the platform is crucial, not only to protect the information of users but also because of the sensitive nature of health data stored on the HDN. Using a public cloud provider also makes it possible to control the region where the data is stored and processed in order to comply with local regulations. Additional controls have been implemented which prevent data from being directly downloaded or removed from a secure Google Cloud bucket. Using a public cloud provider also allows control over the region where the data is stored and processed to comply with local regulations.

We leveraged the cloud to make HDN highly scalable allowing users to access their desired level of computing power. Due to cloud computing, the Health Data Nexus is not bound by hardware resources of the academic institution and can scale on-demand to serve the needs of researchers. Using a cloud platform also makes it easier to control billing for used resources. We developed an architecture that allows for use of external billing methods, vastly simplifying the operational overhead on TCAIREM’s end and allows for a self-service process of creating research environments.

Finally, we developed a streamlined computational infrastructure, allowing for a clear distribution of responsibilities and standardized tools for the reuse of well-established technological stacks and frameworks. For users, the backend provides templates for research environments with the desired configuration, with an ability to launch Jupyter and RStudio-based environments and connect them to cloud services. Other analytics tools and research environments can be configured for access if needed. Upon a user’s request for a new research environment, the backend handles the allocation of necessary cloud resources, the deployment of specified tools, and the authorization to datasets through appropriate Google Cloud Platform policies. It then simplifies dataset access by presenting them through the familiar file system provided by the Cloud Storage FUSE tool. The backend orchestrates administrative actions within the Google Cloud Platform, streamlining the creation of Cloud Identity for users and managing their billing account setups. It supports custom billing arrangements, such as sharing a billing account among multiple users from the same organization.

The flexibility provided by the cloud infrastructure is also important for platform development. New features, types of environments, and tools can be added dynamically as needed. The backend also orchestrates processes like managing data access using cloud infrastructure, provisioning environments, and controlling billing (Supplementary Figures 1 and 2).

### Governance model

The Health Data Nexus is built under the context of Canadian and Ontario privacy legislation and research guidelines, which inform the governance model of the platform. Relevant documents include Ontario’s Personal Health Information Protection Act (PHIPA) (2004) [7] and the Tri-Council Policy Statement on Ethical Conduct for Research Involving Humans (2022) [8]. These provisions correspond with HIPAA (1996) [9] in the United States, whose guidelines have also been followed in the development of platform governance. The governance model of the Health Data Nexus has been reviewed by a Privacy Impact Assessment conducted by a highly specialised legal firm. In addition, the technical capabilities and security have been reviewed by a Threat Risk Assessment.

### Data de-identification

To avoid potential complications related to working with personal health information under PHIPA, the Health Data Nexus exclusively works with de-identified health data. The governance process begins with T-CAIREM signing a Data Transfer Agreement with an interested party (the Data Holder; Figure 2A). The Data Holder is responsible for de-identifying the data to an appropriate standard (as determined by T-CAIREM and informed by the Information and Privacy Commissioner of Ontario [10]) and for providing the dataset to T-CAIREM after any appropriate pre-processing. The Data Holder will also provide associated documentation and metadata needed to understand and use the data and will upload the dataset to the HDN with the help of the T-CAIREM Data Steward.

### User credentials

Before logging into the platform, potential users can view the collection of datasets available on the platform. They can then access the dataset by logging into the platform by Single Sign-on through their university or academic institution. Once logged in, a user can become an Authorized Dataset User for a particular dataset by doing the following (Figure 2B):

1. Becoming Credentialed on the platform by submitting personal information and a reference
2. Completing any required training associated with the dataset (at minimum TCPS 2: CORE-2022 training associated with the Tri-Council Policy Statement on Ethical Conduct for Research Involving Humans) [8]
3. Signing a Data Use Agreement, unique to each dataset and approved by both T-CAIREM and the Data Holder.

An Authorized Dataset User is given full access to the requested dataset, and can access it by creating a Cloud Identity and a Research Environment for the purposes of Analysis.

### Data access levels

All described governance provisions apply to all datasets on the platform; however, data holders may wish to provide additional protection for particularly sensitive datasets. To this end, we have introduced three levels of data access (Figure 2A). The first level of access (‘Zone 1’) includes all requirements outlined above. The second level of access (‘Zone 2’) includes all requirements outlined above, in addition to requiring a research plan to be submitted by the interested user, which must be approved by the data holder. The third and final level of access (‘Zone 3’) includes all requirements outlined above, in addition to requiring a research plan which must be approved not only by the data holder but also by an appropriate Research Ethics Board (REB). Higher zones of access may be appropriate when dealing with potentially sensitive data, or when data holders want community stakeholders to play a role in determining dataset access.

### Comparison with other data platforms

While there exist prominent data platforms, the Health Data Nexus platform displays a unique combination of features (Table 1). This list of platforms is non-exhaustive; other platforms exist in the literature as a point of comparison [11, 12, 13]

**Table 1.** A comparison of the offerings on different data platforms.

| Platform | Available Data | Analysis Options | Security | Accessibility |
| --- | --- | --- | --- | --- |
| Health Data Nexus | Approx. 10 curated and well-documented datasets covering topics related to | Data can be accessed and analyzed in a cloud computing environment after a brief | Data cannot be directly downloaded and is subject to data transfer and data use agreements, | Account registration, credentialing, and training can be completed within a span of hours, |
|  | medicine and health | onboarding process. | allowing for secure sharing of patient data. | giving users same-day access to datasets. |
| Kaggle [14] | Almost 450 000 datasets across a wide variety of fields; datasets can be uploaded by any user with minimal curation | Data can be directly downloaded and analyzed, or uploaded into an online analysis platform | Data security is minimal, and the platform is not suited for hosting potentially sensitive data. | Data can be immediately downloaded and accessed. |
| PhysioNet [2] | Over 350 datasets, curated for the platform, in addition to software and resources. | Data can be downloaded directly or accessed in Google BigQuery | Data is curated and reviewed before hosting, but direct downloads mean data is not protected against re-identification or linkage attacks. | As with Health Data Nexus, most datasets can be accessed with same-day approval. |
| ICES (formerly Institute for Clinical Evaluative Sciences) [15] | Over 100 datasets, primarily related to health services operation | Data is only accessible upon review of a project plan approved by an ICES scientist, available in a limited analysis environment. | ICES is a prescribed entity per PHIPA, meaning it can safely and securely host data (including sensitive data which includes identifiable information) | An in-depth data access form is required, taking time to put together as well as time to review. |

**Table 2.** An overview of the datasets available on the Health Data Nexus.

| Dataset | Abbrev. | Source | Summary |
| --- | --- | --- | --- |
| General Internal Medicine | GIM | St. Michael's Hospital | In-patient data drawn from EHR, Admit Discharge Transfer System, and Medication Administration Check System on 22000 patient encounters |
| Cervical Spine CT Scan | CSpine | St. Michael's Hospital | 1000+ merged CT scans in DICOM format |
| COVID-19 In-Patient Datasets | THP-C19 | Trillium Health Partners | Demographic, clinical, and outcome-related data on 500+ patients |
| Canadian Heart Health Database | CHHDB | Canadian Heart Health Initiative | Country-wide survey data on 23 000 entries across all ten provinces |
| Trial Files | - | Mount Sinai Hospital | Summary of the results of randomized control trials published in clinical journals. |
| Canadian Community Health Survey | CCHS | Statistics Canada | Nationwide survey on health status, healthcare utilization, and health determinants |
| COVID-19 Epidemiology and Vaccination Data | - | COVID-19 Canada Open Data Working Group | Epidemiological dataset for COVID-19 |
| Sleep Laboratory Dataset | - | Sunnybrook Research Institute | Data from Sunnybrook Sleep Laboratory that includes de-identified raw overnight signals, scored sleep metrics, sleep and health questionnaires, and medications/medical history |
| Bridge2AI-Voice Dataset | B2AI | Multi-site (based in University of South Florida) | A dataset of voice recordings and metadata to enable the development, benchmarking, and validation of clinically applicable machine-learning models for diagnosing a wide range of health conditions. |

## CASE STUDIES

Early results demonstrate the capacity of the Health Data Nexus in facilitating data acquisition and educational opportunities.

### Data Acquisition

One of the key value adds of the Health Data Nexus has been providing data storage at no cost. This provides an effective way to engage Data Holders and spread interest in the platform.

As of May 2024, the Health Data Nexus hosts 7 datasets comprising structured tabular data, images, and population-level health data.. The St. Michael’s Hospital General Internal Medicine (GIM) dataset [16] is the flagship dataset for the platform. This dataset provides a wide range of patient information on 14 000 patients over 22 000 visits, including outcomes, tests, medication information, and demographics, and provides a gold standard of tabular data on the platform. Another key dataset is the Cervical Spine CT Scan (CSpine) Dataset, also from St. Michael’s Hospital [14]. This is the largest imaging dataset on the platform, containing a total of over 1000 CT scans. This sets a precedent for HDN hosting a wide variety of datasets of different modalities.

The other datasets on the platform are the COVID in-patient dataset from Trillium Health Partners [17], Canadian Heart Health Database [18], Trial Files for Randomized Control Trials on Large Language Models [19], Canadian Community Health Survey [20], COVID-19 Epidemiology and Vaccination data [21], Sleep Laboratory Dataset [22], and Bridge2AI-Voice Dataset [23].

### User Diversity

HDN attracts a wide range of users from 5 different countries (Fig 4). Users are primarily from Canadian universities in the province of Ontario (93%) with the remaining proportion (7%) being from the United States, Brazil, Poland, the United Kingdom, and the rest of Canada. The growth of the user base over time (Fig 5) reflects the inclusion of new datasets and major events centred on HDN, notably the two Toronto Health Datathons hosted using the platform.

**Figure 4.**
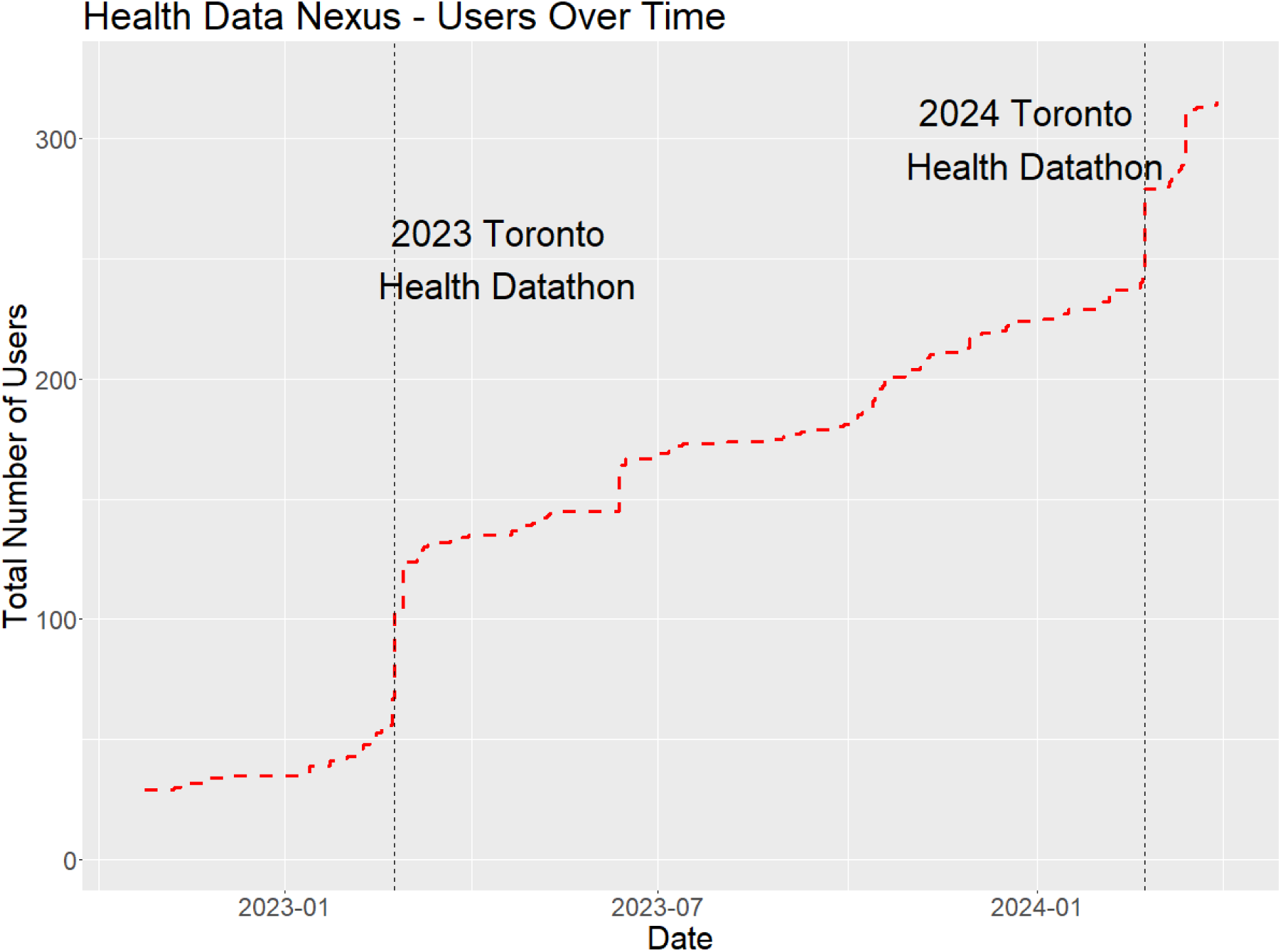
Number of users on the platform over time, highlighting the increase in users corresponding with major events (the 2023 and 2024 Toronto Health Datathons). The GIM dataset was first uploaded on October 21st, 2022, and the CSpine dataset was first uploaded on February 10th, 2023.

**Figure 5.**
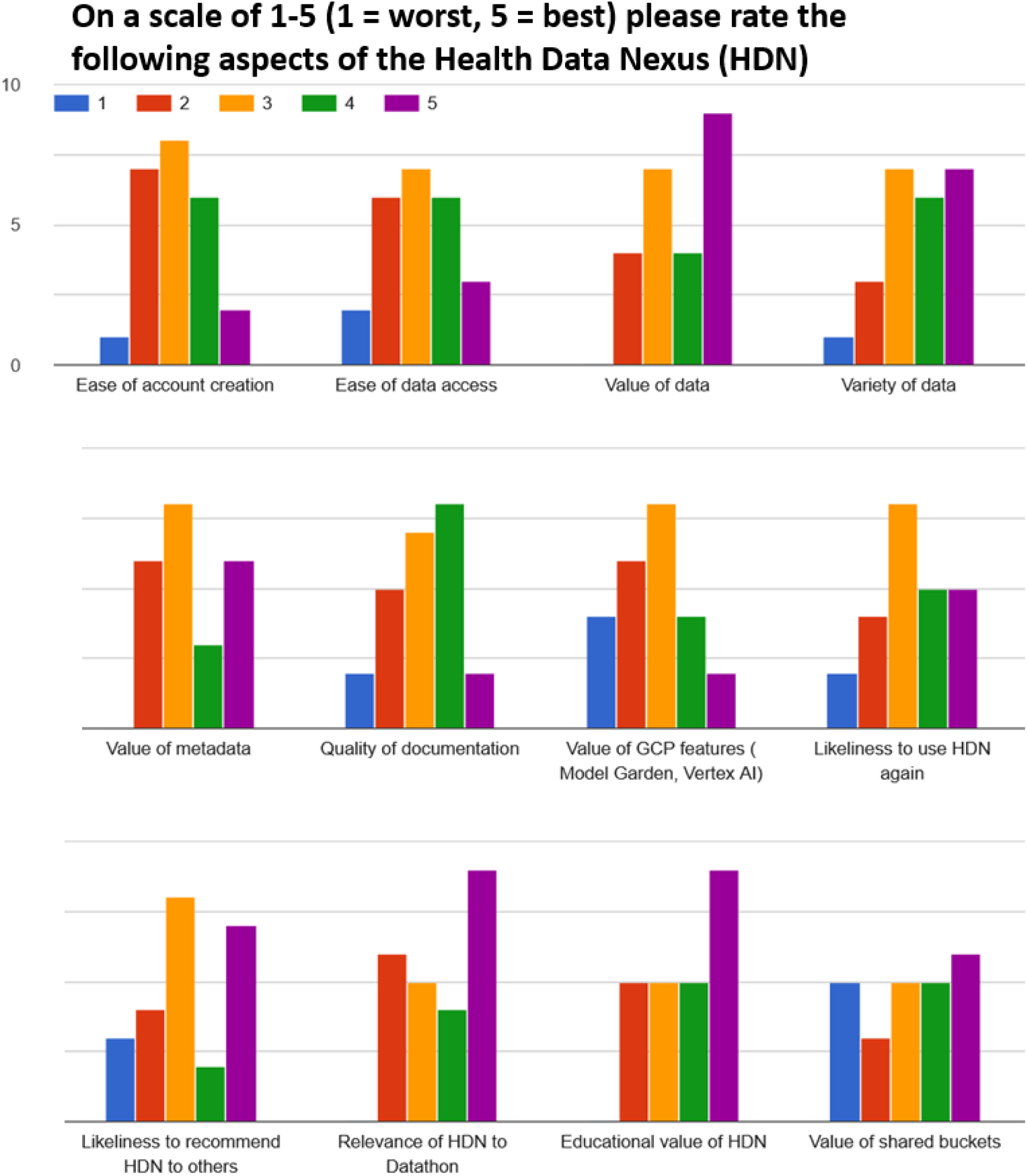
Responses to a post-event survey from the Toronto Health Datathon 2025 on the value of different components to the Health Data Nexus (n=24 responses). Questions included accessibility and value of both the datasets and supplementary features.

### Education

The Health Data Nexus has seen the greatest success as a tool for educators, providing an easy way to provide students and trainees with access to high quality medical datasets. One of the first and ongoing demonstrations of this use case has been through a professor in biomechanical engineering. Over two years, this professor has run short-term projects with students which involve providing access to the GIM [16], COVID-19 [18], and Canada Heart Health [19] datasets and then developing small group projects. Access to real medical data has been helpful for introducing students to some of the challenges of working with real-world data, including inconsistencies, pre-analysis, bias, and class imbalances that are present in real-world health datasets.

Health Data Nexus has also been used for workshops, such as the VADA Summer Schools workshop held between the University of Victoria and the University of Manitoba [25]. This week-long workshop involved a two-day group project for students to get access to datasets and test out analytics. The ease of onboarding has been facilitated through an Events workflow on the platform, which provides a large number of users with an easy way to become credentialed. In addition, the use of shared billing accounts provided by the course administrator or professor dispenses with the requirement of each student using their own billing information, which may be an equity concern. We continue to explore new ways of providing ease of access and providing a fulfilling educational experience for users, such as promoting Google Research Credits [26] for educators and workshop organizers.

### Datathons

The Toronto Health Datathon has been held for three years, in 2023, 2024, and 2025. This event brings students, trainees, data scientists, and healthcare professionals together for a two-day event. Participants join groups to tackle a problem related to one of three available datasets. In both the 2023 and 2024 datathons, datasets have included the General Internal Medicine in-patient tabular data, COVID-19 patient data, and Cervical Spine CT Scan imaging data. In the 2025 datathon, datasets included the General Internal Medicine dataset, the Sleep Study dataset, and the Bridge2AI Voice dataset.

Datathons have provided valuable feedback and suggested improvements to the Health Data Nexus. Between the 2023 and 2024 datathons, improvements to the platforms included providing cloud storage for group members to share code and resources, implementing shared billing accounts, and refining and improving the stability of the platform. In a post-event feedback survey following the 2024 datathon, 19 of 25 participants (76%) agreed or strongly agreed that the Health Data Nexus platform met their expectations. Constructive feedback on the platform included performance issues and providing higher-power computing resources, including access to more powerful GPUs and tools associated with Google’s Vertex AI. This feedback will be incorporated as we continue to develop the platform.

The 2025 Datathon placed an additional focus on evaluation feedback. We have included a summary of the feedback in Figure 5 and highlight a few key results:

- Participants highlighted the value and variety of the data on the platform, including its educational relevance.
- Some participants found that some of the supplemental features of the platform, such as the option to attach a shared bucket to a workspace for sharing code and results, were of limited value in their current state.
- Participants were mixed on the ease of account creation.

## DISCUSSION

### Collaboration between Industry and Academic partners

The Health Data Nexus initiative emerged through a strategic collaboration between academia and industry, combining the resources and expertise of both sectors. Academic partners, including the University of Toronto and Massachusetts Institute of Technology, contributed invaluable domain knowledge, subject expertise, and deep insights into the PhysioNet platform. Conversely, the industry partner played a pivotal role by providing software development expertise, overseeing the development cycle, and implementing best practices in executing technology projects.

Although the collaboration between academic and industry partners – often referred to as Academia-to-Business (A2B) or University-Industry Collaborations (UIC) – holds significant potential for generating valuable results, it is crucial to identify and address common challenges associated with it. Over the course of the project, the team examined, assessed, and adjusted various aspects, including:

1) Understanding of the common vision,
2) Alignment of short- and long-term goals,
3) Effectiveness of collaboration and software development processes,
4) Quality and timely delivery of outcomes.

These areas were identified as major challenges in academic-industry collaborations [27] due to often differing contexts and realities that underlie project motivation (research and curiosity vs. delivery and revenue), planning (discovery, research, open timeline vs. time and budget constraints), and execution in academic and industry partnerships.

To address the challenges commonly encountered, the teams adhered to the principles of Agile Software Development [28]. This methodology underscores the importance of moving towards a common goal through iterative processes, featuring short feedback loops, close stakeholder collaboration, and adaptability to evolving requirements. Embracing this approach proved fundamental throughout the development phase, facilitating prompt and well-informed decision-making, particularly evident in the assessment of various cloud platforms’ capabilities.

Despite the distributed nature of the team, collaboration remained close throughout the project. To ensure timely and frequent feedback the team engaged in week-long development sprints. These sprints incorporated weekly meetings for work review, discussion, and planning, alongside daily synchronous and asynchronous communication to track progress. Throughout the project’s duration, both teams demonstrated responsiveness and active participation, contributing to its success.

Next to making sure the teams can work together towards a goal, an alignment in understanding and approach towards commercial aspects is needed. Just as in any software development project involving multiple stakeholders, it is crucial to establish consensus on critical factors such as Intellectual Property Rights ownership and transfer, commercialization and licensing strategies, resource allocation, risk management protocols, deliverables acceptance criteria, payment agreements, and tracking mechanisms.

### User Curation

An important part of developing the Health Data Nexus as a viable and well-used platform is curating both the selection of datasets and the users on the platform.

It is important to have a wide collection of datasets that cross a variety of subjects and data types. The Health Data Nexus currently includes tabular data of patient records (from both General Internal Medicine and COVID-19 wards) and CT scan imaging data, with plans to expand to include videos, different types of imaging data, genomics data, and de-identified free text, in addition to a wider variety of imaging data. Acquiring these datasets relies on relationship building and demonstrating the value of the Health Data Nexus to interested data holders.

Curating an active and engaged platform user base also relies heavily on building relationships with scientific communities, reaching out via communications and social media, and developing a set of high-value datasets that provide worthwhile value to users. Effective documentation (described below) and streamlined processes for providing support to users are essential for ensuring adoption of the platform.

### Documentation

Provision of useful and accurate documentation for users is a key reason for the success of the Health Data Nexus. Each dataset on the Health Data Nexus is provided with a data page, adapted from the structure used for Physionet, that includes essential information about the dataset. Including background information, methods used in dataset development, usage notes, and a detailed description. This provides users with the essential information needed for their analysis.

Additionally, the Health Data Nexus uses a system of supplemental documentation adapted from the Divio documentation system [29]. This system uses four complementary types of documentation. The Data Dictionary provides detailed information about every field in the data tables included under a dataset. The Explanation and Backgrounds provides additional context on how the dataset was created and for what purpose. The Tutorials provide in-depth guides on performing basic analysis using the dataset. The Getting Started guide connects users with the resources they need to load up the analysis for their first time using the data.

The supplemental documentation is hosted on an external dashboard linked from the data page. The combination of these resources gives users the full set of information they need to work with the data.

Finally, usability is key to ensure that interested researchers, educators, and students have a successful experience with the platform. In addition to the resources provided, a variety of information helps inform new users to the platform, including instructional content on the website and a series of tutorial videos.

### Future Directions

Continued improvement of the Health Data Nexus is a key priority moving forward. Some suggested features include new analysis environments, tools, and research software, and additional computing resources and Google Cloud tools for data analysis. These suggestions reflect the addition of new datasets in a wide variety of fields, including imaging and video analysis, free text data, genomics data, and electrical and physiological signals. In particular, development will include features for sharing relevant code and models among users of a dataset. This will allow for improved educational opportunities among users and the development of a shared body of knowledge. To improve ease of use, development will also include implementing standards such as the Fast Healthcare Interoperability Resources (FHIR) [30] for new and existing datasets on the platform.

Development plans also include involving new partners in the progression of the Health Data Nexus. In addition to attracting new researchers, educators, and data holders, we plan to engage industry. Corporate datathons help upskill those in industry, ensuring they develop and maintain their analysis skills. Incorporating the data collected by private entities also facilitates collaborations between industry and academia to develop new algorithms and pipelines which are relevant to public health.

Finally, we provide the option for other organizations to deploy analogous versions of Health Data Nexus that best suit their local needs, since all code for the platform is open source. Likewise, as the code is open source, we welcome collaboration with other organizations to augment the content and delivery of Health Data Nexus. This is an exciting prospect that could substantially benefit our partners in the future.

## CONCLUSION

We developed the Health Data Nexus platform, and through this article speak to the motivation and design principles through to its development and use by the research community. We highlighted how security, scalability, streamlined design, and a social community of open source developers have led to a successful development, and also how focusing on the education offerings and workshops has spurred interest among users. Ultimately, the Health Data Nexus is positioned to provide a wide variety of multimodal data for educators, trainees, and AI researchers from a variety of backgrounds, with a unique and effective access model. By facilitating access to data, we hope to enrich the medical research ecosystem and aid in the development of patient-centric technologies that will further advance health through AI.

## Data Availability

All data produced are available online at https://healthdatanexus.ai/

## List of Abbreviations

A2B: Academia-to-Business
AI: Artificial Intelligence
BSD: Berkeley Software Distribution
CT: Computed Tomography
FHIR: Fast Healthcare Interoperability Resources
GCP: Google Cloud Platform
GIM: General Internal Medicine
GPU: Graphics Processing Unit
HDN: Health Data Nexus
IRB: Institutional Review Board
ML: Machine Learning
PHIPA: Personal Health Information Protection Act
PIA: Privacy Impact Assessment
REB: Research Ethics Board
T-CAIREM: Temerty Centre for Artificial Intelligence Research and Education in Medicine
TCPS: Tri-Council Policy Statement
TCPS-CORE: Tri-Council Policy Statement - Course on Research Ethics
TRA: Threat Risk Assessment
VADA: Visual and Automated Disease Analytics

## Declarations

### Ethics Approval and Consent to Participate

N/A

### Consent for Publication

N/A

### Data Availability

All data referenced in this publication is available for open access on the Health Data Nexus.

### Competing Interests

The author MM holds non-controlling shares in Signal1 AI. The authors RC, KS, DH, and KR are all employed by Upside Labs.

### Funding

Funding for the creation of T-CAIREM and the Health Data Nexus has been supported by the Temerty Foundation through a transformational gift.

### Authors’ Contributions

JA: Data Curation, Writing - Draft, Writing - Review, Visualization; RC: Conceptualization, Supervision, Project Administration, Resources; KS: Project Administration, Software; DH: Funding, Writing - Review; ZS: Project Administration, Funding; RS: Software, Visualization; MM: Conceptualization, Supervision, Funding, Writing - Review; AJ: Conceptualization, Methodology, Supervision, Software; KR: Project Administration, Resources; TP: Conceptualization, Methodology; DR: Conceptualization, Supervision; BHK: Conceptualization, Supervision, Writing - Draft, Writing - Review.

## Acknowledgements

The authors would like to thank Gemma Postill, Abishek Moturu, and Devin Singh for their feedback and review of the manuscript.

## Notes

### Summary of Updates

New Table 1 includes additional comparative information on data platforms along with context in the text; New Figure 5 includes evaluation data based on use of the platform.

